# Increased Pregnancy Complications Among Mothers with Adverse Childhood Experiences: Findings from a Cross-Sectional Study

**DOI:** 10.1101/2025.09.08.25335335

**Authors:** Mark F.M. Ketelaars, Iris L.M. van Dam, Anne M. de Grauw, Jessica C. Kiefte-de Jong, Mirjam van Veen

## Abstract

**Background:** Adverse Childhood Experiences (ACEs) are potentially traumatic events and family challenges that occur during childhood (0-17 years). Extensive research has linked ACEs to adverse effects throughout life. However, recent studies suggest that ACEs may also have intergenerational consequences, influencing pregnancy complications and perinatal outcomes.

**Objective:** In this study, we aim to explore the association between maternal ACEs and perinatal outcomes in the Dutch context.

**Participants and Setting:** In this retrospective cross-sectional observational study, survey data from 819 mothers with a singleton child under 2 years of age were analysed.

**Methods:** We used logistic and linear regression models to analyse the association between self-reported ACE-10 scores, pregnancy complications and birth outcomes. Adjusted models included maternal education as a proxy for socio-economic position.

**Results:** ACEs were common: 46.3% of mothers had experienced 1-3 ACEs and 17.5% reported ≥4 ACEs. Mothers with ≥4 ACEs had 1.55 times higher odds of pregnancy complications (aOR = 1.55, 95% CI: 1.01–2.37, p = 0.045). Linear regression showed a similar trend (aOR = 1.07, 95% CI: 0.99–1.16, p = 0.085). No associations were found for prematurity, small for gestational age, NICU admission, birth weight, or gestational age.

**Conclusions:** Maternal ACEs were associated with pregnancy complications (gestational hypertension, gestational diabetes, preeclampsia or premature rupture of membranes) but not directly with birth outcomes like prematurity or low birth weight. These findings highlight the importance of early psychosocial risk detection. Strengthening preventive care with systems may help mitigate intergenerational adversity, even without formal ACE screening.

## Introduction

Adverse Childhood Experiences (ACEs) are potentially traumatic events and family challenges that occur during childhood (0-17 years). They are categorized in abuse (physical, emotional and sexual), neglect (physical and emotional) and household dysfunction (substance abuse, mental illness, domestic violence, incarceration and parental separation) (Dube et al., 2003). Adverse childhood experiences are common, with 63.9% of Americans reporting at least one ACE and 17.3% having experienced four or more. In the United Kingdom, ACE exposure is slightly lower but still prevalent: 46.9% of the population has experienced at least one, while 12.6% reported four or more (Hughes et al., 2020; Swedo et al., 2023). While research on ACEs in the Netherlands is scarce compared to the international literature, prevalence rates in the Dutch population appear to be similar to those in the United Kingdom and other parts of Europe. Numbers of Dutch adults experiencing at least one ACE range from 43,9% to 48,7%, with number of four or more ACEs ranging between 5.72% and 11.2% (Bussemakers et al., 2019; Kuiper et al., 2010).

Extensive research spanning three decades has shown that ACEs are consistently associated with adverse effects throughout life. For example, exposure to ACEs is associated with psychological effects such as internalizing and externalizing behaviour problems but also increased risky health behaviour and higher rates of delinquency among adolescents (Dube et al., 2003; Felitti et al., 1998). Moreover, ACEs are associated with higher rates of posttraumatic stress syndrome, diabetes, cancer and cardiovascular diseases – all in a dose-response relationship. Ultimately, an accumulation of ACEs has been found to be associated with many of the leading causes of death, and even premature death itself (Felitti et al., 1998; Neves et al., 2021).

Recently, more and more studies have been suggesting that ACEs may also have intergenerational consequences, hereby affecting pregnancy complications, perinatal outcomes and child development (Miller et al., 2021). Additionally, multiple studies have indicated a relationship between maternal ACEs and preterm birth (Christiaens et al., 2015; Mamun et al., 2023; Sulaiman et al., 2021). There is also a growing body of literature providing evidence that women with higher ACE scores may be at higher risk of perinatal complications compared to women without exposure to abuse. These complications include hypertensive disorders during pregnancy, gestational diabetes mellitus, antenatal depression, and low offspring birthweight (Bublitz et al., 2020; Mamun et al., 2023).

This suggests adverse childhood experiences may alter life trajectories across multiple generations in different ways. Given the importance of the first 1000 days in early development, children born to parents that have experienced ACEs may face lifelong consequences due to an increased risk of these perinatal complications. Hence, primary prevention of ACEs and mitigation of their effects are essential components of an equity-focused public health strategy.

Most studies on the intergenerational effects of ACEs have been conducted in the context of limited universal preventive healthcare systems in early life. However, the Netherlands’ unique preventive youth health system (JGZ) provides universal, non-medical care focused on early detection and psychosocial support both during pregnancy and during childhood, contrasting with more medically oriented systems in other European countries. Moreover, the Dutch Neonatal Follow-Up (LNF) monitors all children born after a complicated perinatal period until at least the age of 8 years. Traditionally, in other European countries, such follow-up programs are generally only designed for study purposes. Within this system, it may be expected that the intergenerational impact of ACEs could be different in the context of The Netherlands but this has not been evaluated yet.

The aim of this study is therefore to explore the prevalence of ACEs in parents of young children in the Netherlands, and to analyse the association of those childhood experiences with perinatal outcomes. We hypothesized that higher maternal ACEs scores are associated with a higher incidence of adverse perinatal outcomes.

## Methods

### STUDY DESIGN AND PARTICIPANTS

We performed a retrospective cross-sectional observational study. Subjects were included between August 2023 and November 2024. The study was conducted in the Netherlands. The appropriate approval that the study was not subject to the Medical Examination Act was granted by the Medical Ethical Committee of the Leiden University Medical Center (LUMC) under reference number 23-011. The study protocol was approved by the Scientific Review Board of the Haga Teaching Hospital in the Hague. Data was collected using online surveys that were initially distributed in a paediatric hospital and children’s health clinics throughout The Hague, where members of the study team handed out flyers to parents. Also, an advert was launched nationwide using Meta Ads, targeting parents on Facebook and Instagram. Parents were eligible to participate if they had a singleton child under the age of 2 years old and sufficient knowledge of the Dutch or English language. A total of 871 respondents completed the survey. The vast majority of the participants (N=822) was the biological mother of their last child. Given the low number of fathers and non-biological mothers, only biological mothers were included in the analysis. The final study population was limited to mothers with available data on ACE exposure and at least one perinatal outcome (N = 819).

### EXPOSURES: ACES

All participants were provided a digital anonymous survey that included the ACE questions according to the Dutch Questionnaire for ACEs in adults and a translated version of the Benevolent Childhood Experiences scale (Kuiper et al., 2010; Narayan et al., 2018). The questionnaire for ACEs assesses 10 different types of adverse experiences: emotional abuse, physical abuse, sexual abuse, emotional neglect, physical neglect, parental separation or divorce, witness to intimate partner violence, household substance abuse, household mental illness and incarcerated household member. Kuiper et al. reported a high internal validity of the ACE questionnaire in the Dutch population (2010). ACEs were scored on a scale of 0-10 and subsequently categorized in three groups as in previous research: 0, 1-3 and ≥4 (Miller et al., 2021). The BCE scale was translated to Dutch by IvD and subsequently back translated to English by MK. Any discrepancies in translation were resolved by mutual agreement and the scientific review board of the Haga Teaching Hospital approved the questionnaire.

### PERINATAL OUTCOMES

Several perinatal outcomes were collected and can be divided in pregnancy complications and birth outcomes. Mothers were dichotomized into groups based on the presence of gestational hypertension, gestational diabetes, preeclampsia and premature rupture of membranes during the last pregnancy. These groups were pooled and combined to create a binary variable for pregnancy complications (yes/no). Birth outcomes included pregnancy duration, birthweight, being small for gestational age (SGA) and whether the child had been admitted to the NICU after birth. Pregnancy duration and birthweight were treated as continuous variables. Pregnancy duration was dichotomized using the cut-off of 37 weeks to create a variable for preterm birth. SGA was calculated using sex, birthweight and pregnancy duration using publicly available reference data for the growth of Dutch children (Bocca-Tjeertes et al., 2012).

### COVARIATES

The collected demographic characteristics included child sex, maternal age (continuous, in years), parental relationship (living together or living apart), postal code (to approximate socioeconomic position (SEP)), religious beliefs (categorical), educational attainment (categorized according to the Dutch education system, then classified as low/middle and higher education), workforce status (full time employed, part time employed, self-employed, other), total yearly household income (ordinal categories with steps of €10.000, then classified as below or above modal), country of origin of both the subject and their parents (self-reported, classified into Dutch-born vs. foreign-born), parity (continuous, then classified as first child or not), previous miscarriages, and previous preterm childbirth (binary).

Lifestyle factors included maternal smoking (never, before pregnancy, during pregnancy, after pregnancy), alcohol consumption (never, before pregnancy, during pregnancy, after pregnancy), and illicit drug use (never, before pregnancy, during pregnancy, after pregnancy). We also inquired about current medication use (open-ended response) and mental health history, including any diagnosis of a psychiatric disorder (yes/no) and history of psychological or psychiatric treatment (yes/no). Positive childhood experiences were assessed using a 10 item Benevolent Childhood Experiences (BCEs) scale (Narayan et al., 2018). This includes 10 positive childhood experiences with promising cultural validity, including supportive relationships with childhood caregivers, friends, teachers, neighbours, and mentors; positive beliefs for coping and self-esteem; enjoyment of school and home life; and predictable home routines. The BCE scale showed strong predictive validity in American population (Narayan et al., 2021). To our knowledge, it has not been used in the Dutch context. Similar to the ACEs, BCEs were scored on a scale of 0-10 and subsequently categorized in three groups (0-5, 6-9 and 10).

### STATISTICAL ANALYSIS

After data cleaning, missing data patterns were inspected to identify any irregularities or unexpected distributions. Three variables had a missing rate exceeding 5%. Due to a logistical error, information on child’s sex was not collected in the first 116 surveys (14.2%). These data were therefore assumed to be missing completely at random. Because child’s sex was required to construct one of the outcome variables, and imputation of outcome data is generally discouraged, these cases were excluded from analyses involving sex-specific outcomes. Since child’s sex is required to determine SGA, the composite variable of any perinatal outcome was consequently unavailable for 11.1% of participants. The total ACE score was missing in 7.0% of participants because respondents could select the option “prefer not to answer”.

A data check for potential outliers revealed one extreme outlier in birthweight (350 grams at 40 weeks of pregnancy), which was excluded. Additionally, ten cases of macrosomia (>4500 grams birthweight) were identified, but not excluded because the values were plausible and the total percentage of macrosomia in the sample (1.21%) did not exceed national prevalence rates. Categorical and composite variables were checked and found to be consistent.

To analyse the association between ACEs and perinatal outcomes, logistic and linear regression models were used. Separate univariate models were run for each outcome (prematurity, SGA, pregnancy complications, NICU admission, any perinatal outcome, birthweight and gestational age) with ACEs (both analysed categorically and continuously). Potential confounders were selected a priori based on literature findings and a directed acyclic graph (DAG; see Supplementary Figure 1). In addition, we applied a change-in-estimate (CIE) approach, adding each variable individually to the univariate model for ‘any perinatal outcome’. A variable was retained if the exposure coefficient changed by ≥10% (Lee, 2014). Based on this criterion, maternal education was identified as a confounder prior to analysis and was included consistently in all adjusted models, regardless of statistical significance or model fit. Statistical tests of hypotheses were two-tailed with significance set at *p*<0.05. All statistical analyses were carried out using R (version 4.1.2; R Development Core Team). ChatGPT (OpenAI, 2024) was used to revise parts of the manuscript text. The authors reviewed and verified all content.

## Results

Exposure and outcome data were complete for a total of 819 participants. Table 1 shows the demographic characteristics of the participants stratified by ACE exposure group. The mean maternal age ranged from 31.13 to 31.72 years across the ACE groups. The vast majority of respondents (ranging from 93.0% to 98.3% across groups) reported living with their partner. Participants with high ACE exposure were somewhat less likely to be living together compared to those with no ACEs (7.0% vs. 1.7%). Most participants (72.6% to 78.6%) reported having no religious beliefs, and the majority had attained higher education (71.3% to 89.6% across groups). Participants with high ACE exposure were more likely to report lower educational attainment relative to those with no ACEs (28.7% vs. 10.4%). In terms of employment, 50.3% to 64.0% of participants were employed part-time, while 20.3% to 26.3% were employed full-time, and 5.1% to 20.3% reported being unemployed, or in another category (e.g., student, receiving benefits). Participants with high ACE exposure were more likely to fall into the “other” employment category compared to those with no ACEs (20.3% vs. 5.1%). Household income was reported as above modal income by 30.8% to 44.8% of participants. Participants with high ACE exposure more frequently reported a below modal household income compared to participants with no ACEs (69.2% vs. 55.2%). Regarding reproductive history, 42.7% to 53.5% of participants were experiencing their first pregnancy, and a history of miscarriage was reported by 25.3% to 33.6%.

**Table 1.** Sociodemographic characteristics of participants and summary of variables (N = 819 participants).

| Variables | ACE Score group |  |  | p-value |
| --- | --- | --- | --- | --- |
|  | High<br>N = 143 | Moderate<br>N = 379 | None<br>N = 297 |  |
| BCE Score |  |  |  | <0.001 |
| High | 3 (2.1) | 42 (11.1) | 42 (14.1) |  |
| Moderate | 85 (59.4) | 296 (78.1) | 250 (84.2) |  |
| Low | 55 (38.5) | 41 (10.8) | 5 (1.7) |  |
| Maternal age, mean (SD) | 31.13 (3.29) | 31.57 (3.23) | 31.72 (2.80) | 0.175 |
| Parental relationship |  |  |  | 0.018 |
| Living together | 133 (93.0) | 361 (95.3) | 292 (98.3) |  |
| Not living together | 10 (7.0) | 18 (4.7) | 5 (1.7) |  |
| Religion |  |  |  | 0.341 |
| Religious | 37 (27.4) | 80 (21.4) | 72 (24.4) |  |
| Not religious | 98 (72.6) | 293 (78.6) | 223 (75.6) |  |
| Education |  |  |  |  |
| Higher education | 102 (71.3) | 320 (84.4) | 266 (89.6) | <0.001 |
| Lower/middle education | 41 (28.7) | 59 (15.6) | 31 (10.4) |  |
| Employment |  |  |  | <0.001 |
| Fulltime employed | 29 (20.3) | 99 (26.1) | 78 (26.3) |  |
| Part-time employed | 72 (50.3) | 231 (60.9) | 190 (64.0) |  |
| Self-employed | 13 (9.1) | 19 (5.0) | 14 (4.7) |  |
| Other | 29 (20.3) | 30 (7.9) | 15 (5.1) |  |
| Income |  |  |  | 0.017 |
| Above modal | 44 (30.8) | 160 (42.2) | 133 (44.8) |  |
| Below modal | 99 (69.2) | 219 (57.8) | 164 (55.2) |  |
| Maternal country of birth |  |  |  | 0.093 |
| Netherlands | 132 (92.3) | 359 (94.7) | 288 (97.0) |  |
| Other | 11 (7.7) | 20 (5.3) | 9 (3.0) |  |
| Grandmother's country of birth |  |  |  | <0.001 |
| Netherlands | 116 (81.1) | 334 (88.1) | 278 (93.6) |  |
| Other | 27 (18.9) | 45 (11.9) | 19 (6.4) |  |
| Grandfather's country of birth |  |  |  | 0.041 |
| Netherlands | 122 (85.3) | 340 (89.7) | 276 (92.9) |  |
| Other | 21 (14.7) | 39 (10.3) | 21 (7.1) |  |
| Previous pregnancy |  |  |  | 0.018 |
| No prior pregnancy | 61 (42.7) | 214 (56.5) | 159 (53.5) |  |
| Prior pregnancy | 82 (57.3) | 165 (43.5) | 138 (46.5) |  |
| Previous miscarriage |  |  |  | 0.147 |
| No | 95 (66.4) | 283 (74.7) | 220 (74.1) |  |
| Yes | 48 (33.6) | 96 (25.3) | 77 (25.9) |  |
| Previous preterm birth |  |  |  | 0.673 |
| No | 132 (93.0) | 359 (94.7) | 281 (94.9) |  |
| Yes | 10 (7.0) | 20 (5.3) | 15 (5.1) |  |

Previous preterm birth was reported by 5.1% to 7.0%, which is in line with national figures in the Netherlands (Perined, 2022). Exposure to ACEs was common among participants: 36.3% of the sample reported no ACEs, 46.3% had experienced between one and three ACEs (moderate exposure), and 17.5% reported high ACE exposure (≥4 ACEs). BCEs were reported at varying levels: 12.3% of participants had low BCE scores (0–5), 77.0% fell into the moderate category (6–9), and 10.6% reported the maximum BCE score (10). Participants with high ACE exposure were also more likely to report low BCE scores (38.5% vs. 1.7%).

Table 2 shows unadjusted perinatal outcomes by ACE category. Mean pregnancy duration was 39 weeks across all groups and mean birthweight was 3411 grams. Absolute number of cases for very preterm birth was low in all groups. Gestational age, birthweight, preterm birth, SGA, and NICU admission did not differ significantly between groups. Pregnancy complications were more common in the high ACE group (39.9%) compared to the moderate (31.7%) and no ACE groups (28.3%, p = 0.050). The composite outcome occurred in 51.1% of the high ACE group versus 41.3% in the other groups (p = 0.108).

**Table 2.**
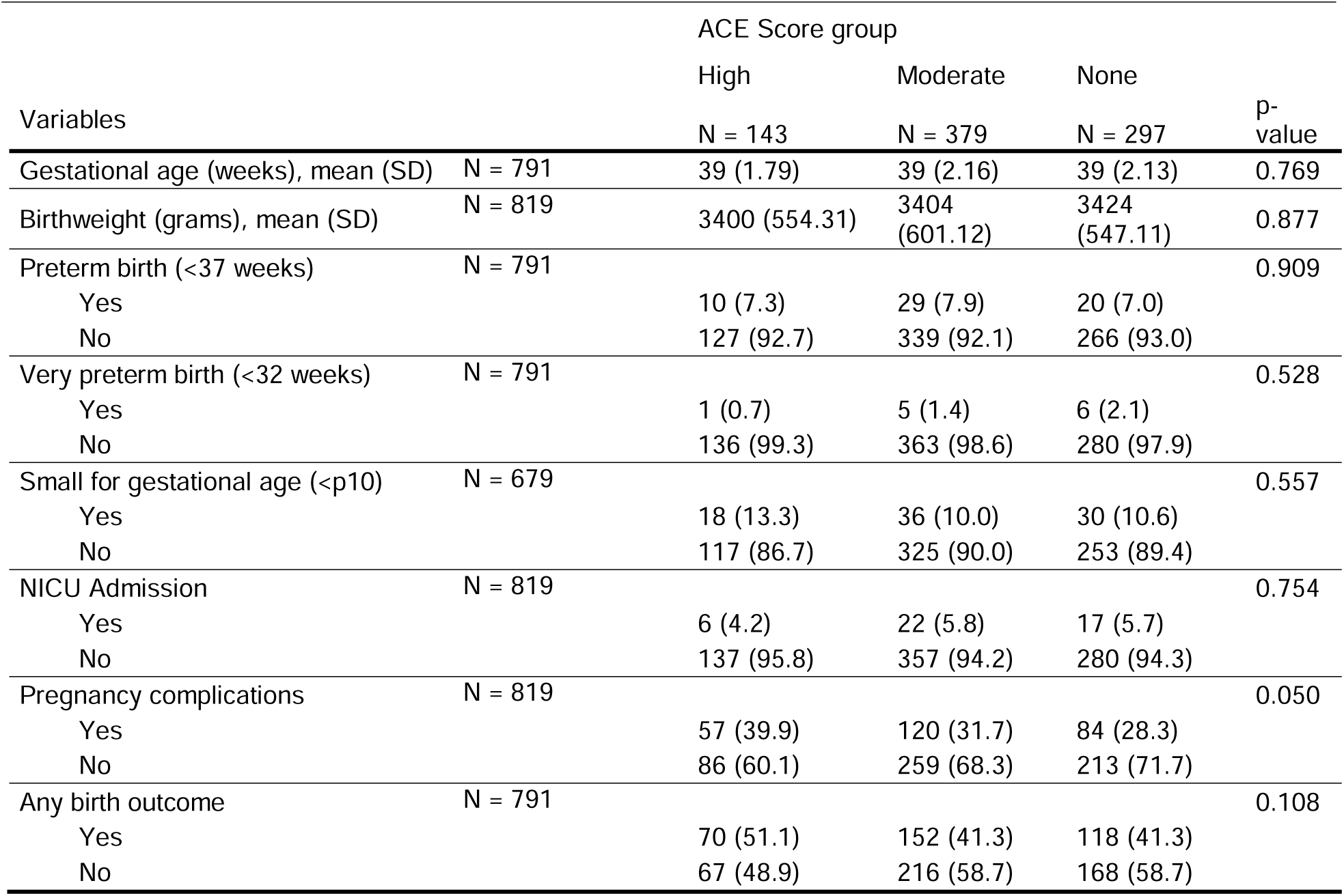
Unadjusted outcomes by ACE category.

The results of the logistic regression models examining categorical ACEs and perinatal outcomes are presented in Table 3. These analyses showed that higher ACE exposure was significantly associated with pregnancy complications. This association remained after adjustment for maternal education. Compared to mothers with no history of ACEs, mothers who reported ≥4 ACEs had 1.55 times higher odds of pregnancy complications (aOR = 1.55, 95% CI: 1.01–2.37, p = 0.045). A trend was observed for the composite perinatal outcome in the unadjusted model, but this association did not remain significant after adjustment for maternal education (aOR = 1.35, 95% CI: 0.89–2.06, p = 0.155). ACES were not significantly associated with preterm birth and SGA.

**Table 3.** Results of the logistic regression between ACE categories and different perinatal outcomes.

| Variable | Category | Unadjusted model |  | Adjusted model |  |
| --- | --- | --- | --- | --- | --- |
|  |  | Estimate (95% CI)<sup>1</sup> | p-value | Estimate (95% CI)<sup>1</sup> | p-value |
| Prematurity | Moderate | OR 1.14 (0.63–2.06) | 0.669 | aOR 1.11 (0.61–2.01) | 0.737 |
|  | High | OR 1.05 (0.48–2.30) | 0.909 | aOR 0.95 (0.43–2.12) | 0.903 |
| SGA | Moderate | OR 0.93 (0.56–1.56) | 0.794 | aOR 0.93 (0.56–1.56) | 0.786 |
|  | High | OR 1.3 (0.70–2.42) | 0.413 | aOR 1.28 (0.68–2.42) | 0.441 |
| NICU Admission | Moderate | OR 1.01 (0.53–1.95) | 0.964 | aOR 1.01 (0.53–1.95) | 0.966 |
|  | High | OR 0.72 (0.28–1.87) | 0.502 | aOR 0.72 (0.27–1.89) | 0.504 |
| Pregnancy complications | Moderate | OR 1.17 (0.84–1.64) | 0.342 | aOR 1.15 (0.82–1.60) | 0.422 |
|  | High | **OR 1.68 (1.11–2.56)** | **0.015 \*** | **aOR 1.55 (1.01–2.37)** | **0.045 \*** |
| Any perinatal outcome | Moderate | OR 1.00 (0.73–1.37) | 0.991 | aOR 0.97 (0.71–1.33) | 0.860 |
|  | High | OR 1.49 (0.99–2.24) | 0.057 | aOR 1.35 (0.89–2.06) | 0.155 |
| Birthweight (grams) | Moderate | $\beta$ -19.9 (-107.1–62.3) | 0.654 | $\beta$ -16.48 (-103.8–70.9) | 0.712 |
| | High | $\beta$ -24.4 (-138.9–90.1) | 0.676 | $\beta$ -12.16 (-128.3–104.0) | 0.837 |
| Gestational age (weeks) | Moderate | $\beta$ -0.09 (-0.41–0.23) | 0.578 | $\beta$ -0.07 (-0.39–0.25) | 0.250 |
| | High | $\beta$ 0.04 (-0.39–0.47) | 0.856 | $\beta$ 0.12 (-0.31–0.55) | 0.577 |

Reference group: no ACEs. Adjusted models include maternal education as covariate.
<sup>1</sup> Logistic outcomes are reported as odds ratios (OR); continuous outcomes as regression coefficients ( $\beta$ ).
\* indicates statistical significance ( $p < 0.05$ ).

Logistic regression models with continuous ACE scores showed similar results, with a significant association for pregnancy complications and a trend for any perinatal outcome (Table 4). However, the association with pregnancy complications did not remain significant after adjustment for maternal education (Table 4). In linear regression models (Tables 3 and 4), no associations were found between ACEs (whether analysed as categories or as a continuous score) and birthweight or gestational age. All complete models, including effect sizes for covariates can be found in Online Supplementary Appendix 1.

**Table 4.** Results of the logistic and linear regression between continuous ACEs and different perinatal outcomes.

| Predictor | Unadjusted model |  | Adjusted model |  |
| --- | --- | --- | --- | --- |
|  | 95% CI | p-value | 95% CI | p-value |
| Prematurity | OR 1.04 (0.91–1.19) | 0.545 | aOR 1.02 (0.89–1.18) | 0.745 |
| SGA | OR 1.03 (0.91–1.16) | 0.633 | aOR 1.02 (0.91–1.16) | 0.704 |
| NICU Admission | OR 0.94 (0.79–1.12) | 0.490 | aOR 0.95 (0.80–1.13) | 0.581 |
| Pregnancy complications | **OR 1.09 (1.01–1.17)** | **0.030 \*** | aOR 1.07 (0.99–1.16) | 0.085 |
| Any perinatal outcome | OR 1.07 (0.99–1.15) | 0.088 | aOR 1.05 (0.97–1.13) | 0.226 |
| Birthweight (grams) | $\beta$ -10.7 (-31.8–10.5) | 0.324 | $\beta$ -8.54 (-30.2–13.2) | 0.440 |
| Gestational age (weeks) | $\beta$ 0.00 (-0.04–0.08) | 0.937 | $\beta$ 0.01 (-0.07–0.09) | 0.775 |

Adjusted models include maternal education as covariate.
<sup>1</sup>□ Logistic outcomes are reported as odds ratios (OR); continuous outcomes as regression coefficients ( $\beta$ ).
\* indicates statistical significance ( $p < 0.05$ ).

## Conclusions

In this study, we showed that maternal ACEs were associated with increased odds of pregnancy complications (gestational hypertension, gestational diabetes, preeclampsia and premature rupture of membranes) whereas no associations were found for other birth- and pregnancy outcomes.

These findings are partially in line with previous research. A meta-analysis by Mamun et al. provides the most comprehensive overview of associations between maternal ACEs and pregnancy complications and birth outcomes to date, reporting a combined “moderate association” (OR 1.39, 95% CI: 1.11–1.74) (2023). For gestational diabetes, five out of six studies in this meta-analysis on gestational diabetes described an association with ACEs, supporting our findings. To our knowledge, only one additional study has been published since the publication of this meta-analysis. Lovett et al. reported that women in the United States with high levels of early life trauma had an increased risk of gestational diabetes, while Swedo et al. found no association in their sample covering five States (Lovett et al., 2024; Swedo et al., 2023). For pregnancy hypertension, evidence remains limited. Only one study was included in the meta-analysis, and it found no association in a sample of a Hispanic community in the United States. Since then, Lovett et al. reported a positive association with gestational hypertension, whereas Swedo et al. again found no significant relationship. Our findings provide more support for a relationship between maternal ACEs and gestational hypertension, gestational diabetes, preeclampsia and premature rupture of membranes.

In the aforementioned meta-analysis, ACEs were associated with preterm birth and low birthweight. Mamun et al. reported combined odds ratios of 1.41 (95% CI 1.16 to 1.71) and 1.27 (95% CI 1.17 to 1.47), respectively (2023). Contrary to the results of this meta-analysis, Sosnowski et al. found no direct relationship between maternal childhood adversity and infant birthweight, gestational age, and NICU admission (2023). However, higher odds of NICU admission were reported by Ciciolla et al. (2021). Overall, the evidence in the literature is contradictive.

The differences in results of this study and some of the findings in the literature could also be explained by the context in which most studies were conducted. To our knowledge, this is the first study on this subject in the Netherlands and only one of the first in European context. The vast majority of studies investigating ACEs and perinatal outcomes was conducted in the United States, where access to (prenatal) healthcare is less universal. In this field of research, context can be of crucial importance. For example, Bronstein et al. highlighted that the maternal health status in the United States generally is poorer compared to many European countries, and unintended pregnancies more common. Because of this, the country scores lower on many perinatal indicators (Bronstein et al., 2018). The overrepresentation of American studies can limit the generalizability of findings to countries with different social, healthcare, and reproductive policy environments. More specifically, the Dutch Solid Start program was initiated nationwide in the Netherlands in 2018. This program offers support to (future) parents that start preconception (Molenaar et al., 2023). Enabling parents to be better prepared for pregnancy may mitigate adverse effects of ACEs on birth complications.

Furthermore, contradicting findings may be explained by the possibility that ACEs exert their effects in an indirect way. For example, the pathway from early life adversity to perinatal outcomes may be mediated by maternal health during pregnancy, rather than showing as a direct effect. We were not able to take such mediating factors into account in our study. Moreover, there may be additional pathways between childhood adversity and perinatal adversity that were not captured in our analyses. For instance, preterm birth is a heterogeneous outcome with diverse biomedical causes, including infections, placental abnormalities, and hypertensive disorders of pregnancy; for example, a recent meta-analysis found that maternal influenza infection increased the odds of preterm birth by 52% (OR = 1.52, 95% CI: 1.18–1.90) (Wang et al., 2023). These factors may overshadow or have a larger effect than psychosocial risk mechanisms or mask associations with ACEs. Also, we were unable to distinguish between spontaneous and medically indicated preterm birth, limiting our ability to assess specific underlying pathways.

Lastly, the prevalence of ACEs among mothers in this study was higher than numbers previously reported, which may influence the results of our analysis. In 2010, Kuiper et al. found that 14% of Dutch women reported having experienced ≥4 ACEs, and 34% reported having 1-3 ACEs. In our study, these levels rose to 18% and 45%, respectively (Kuiper et al., 2010). This increase can partly be explained by higher numbers of divorces in the Netherlands (Statistics Netherlands, 2024). It is, however, unlikely that divorce alone is the driver behind the increase. The rise in both the moderate (1-3) and high (≥4) ACE group implies that other adversities (e.g. household mental illness, substance use, emotional neglect) may also have become more common – or at least more frequently reported, possibly due to more awareness. In the Dutch context, with a system focused on early detection, psychosocial support, and early intervention, the impact of early life adversity may be more effectively mitigated. This could partly explain why some associations found in previous studies, particularly in U.S. settings, were not observed in our study.

In order to appreciate the results, some limitations need to be acknowledged. First, despite efforts to distribute the questionnaire as widely as possible, our sample consisted of mainly highly educated mothers (84.0%) and likely subject to response bias. Second, outcomes like prematurity and SGA are relatively uncommon in the Dutch population (∼10%). The percentage of mothers that had experienced one or more pregnancy complications was much higher (41.5%). Hence, with our sample size, it is plausible that our statistical power was sufficient to detect associations with pregnancy complications, but too small for the less frequently observed outcomes. Third, the traditional ten items in the ACE-10-index have several shortcomings. For example, despite their strong associations with health outcomes, these ten items do not include important adversities such as poverty, community violence, and peer victimization. Moreover, several items are oversimplified, and the binary response format omits certain nuances (McLennan et al., 2020). This makes the ACE-10 suboptimal for clinical use. Nevertheless, given the relevant set of core experiences and the widespread use of the questionnaire, ACEs might be preferred in public health research (Remmers et al., 2024).

Lastly, differences in neonatal care terminology may have affected mothers’ responses regarding NICU admission. Our questionnaire specifically asked about NICU admission, whereas in the Netherlands, neonatal intensive care is centralized in a limited number of teaching hospitals. Many preterm infants, especially those born between 32 and 36 weeks, are routinely admitted to specialized neonatal care units in general hospitals, which may not be labelled as ‘NICU’ by healthcare staff or perceived as such by parents. As a result, both under- and over reporting of NICU admissions may have occurred, even among infants born before 32 weeks of gestation.

Maternal ACEs were associated with increased odds of pregnancy complications, including gestational hypertension, diabetes, preeclampsia, and premature rupture of membranes. No direct association was found with preterm birth, small for gestational age, or low birthweight, despite mixed evidence in the literature. With ACE prevalence in Dutch women rising, monitoring their impact on perinatal health remains critical. The Dutch preventive youth healthcare system is unique in its universal, accessible, and non-medical approach to early detection and psychosocial support. Unlike in countries where perinatal and youth care is primarily delivered through general practitioners or paediatricians, the JGZ offers routine monitoring of maternal and child wellbeing for nearly all families. Our findings underscore the relevance of addressing maternal ACEs within this preventive framework, as early identification of psychosocial risk factors may help reduce intergenerational adversity. Despite the Dutch system, ACEs and their long-term consequences may still be under recognized in standard practice. Although clinical screening and recognition remain a challenge, using trauma informed care and further strengthening antenatal care pathways, mental health support, and social safety nets may reduce the long-term impact of ACEs, even in the absence of formal screening.

## Data Availability

All data produced in the present study are available upon reasonable request to the authors

## Contributions

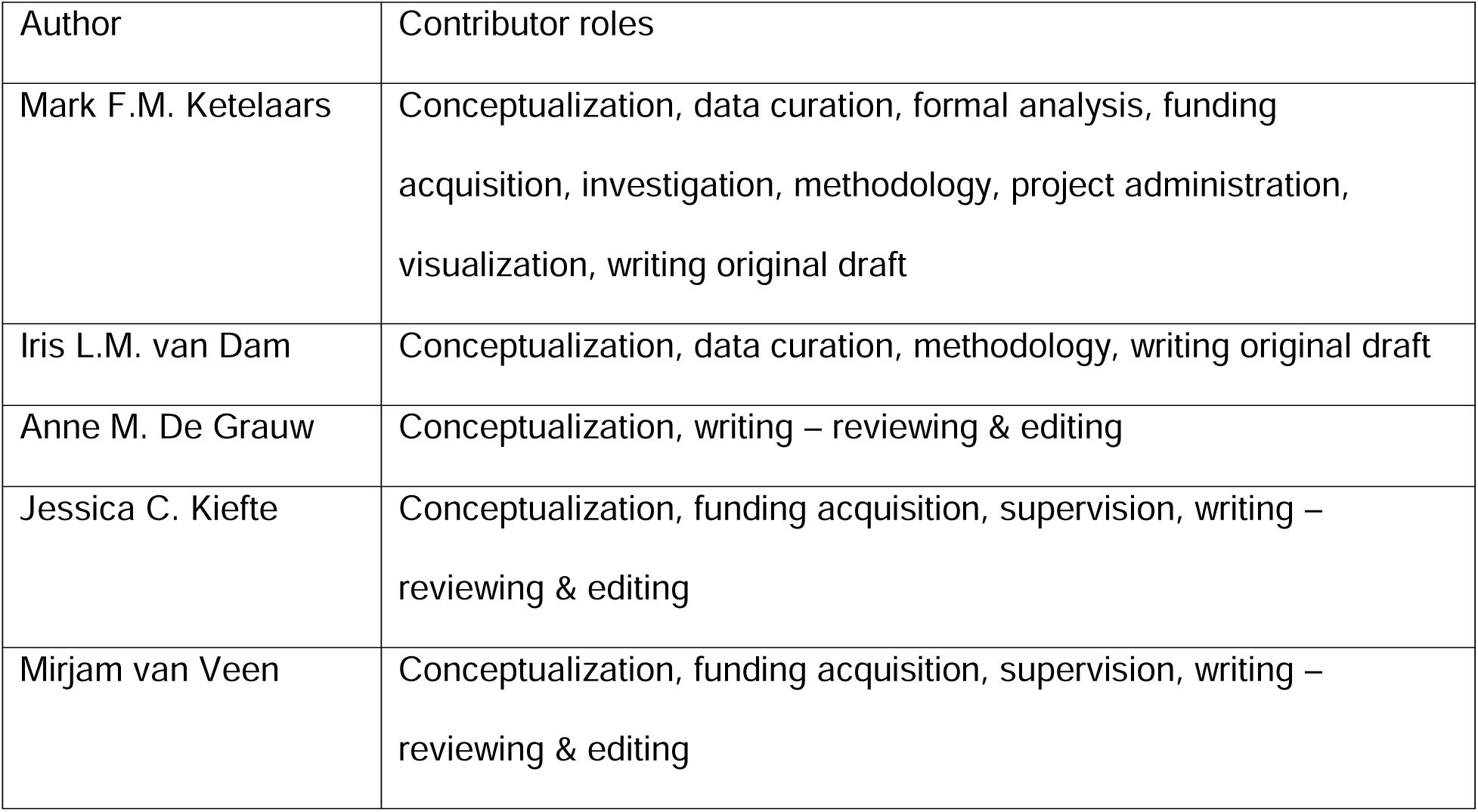

