## Supplementary Figure for "Increased Pregnancy Complications Among Mothers with Adverse Childhood Experiences: Findings from a Cross-Sectional Study"

**Supplementary Figure 1. Directed acyclic graph for covariates and confounder selection.**

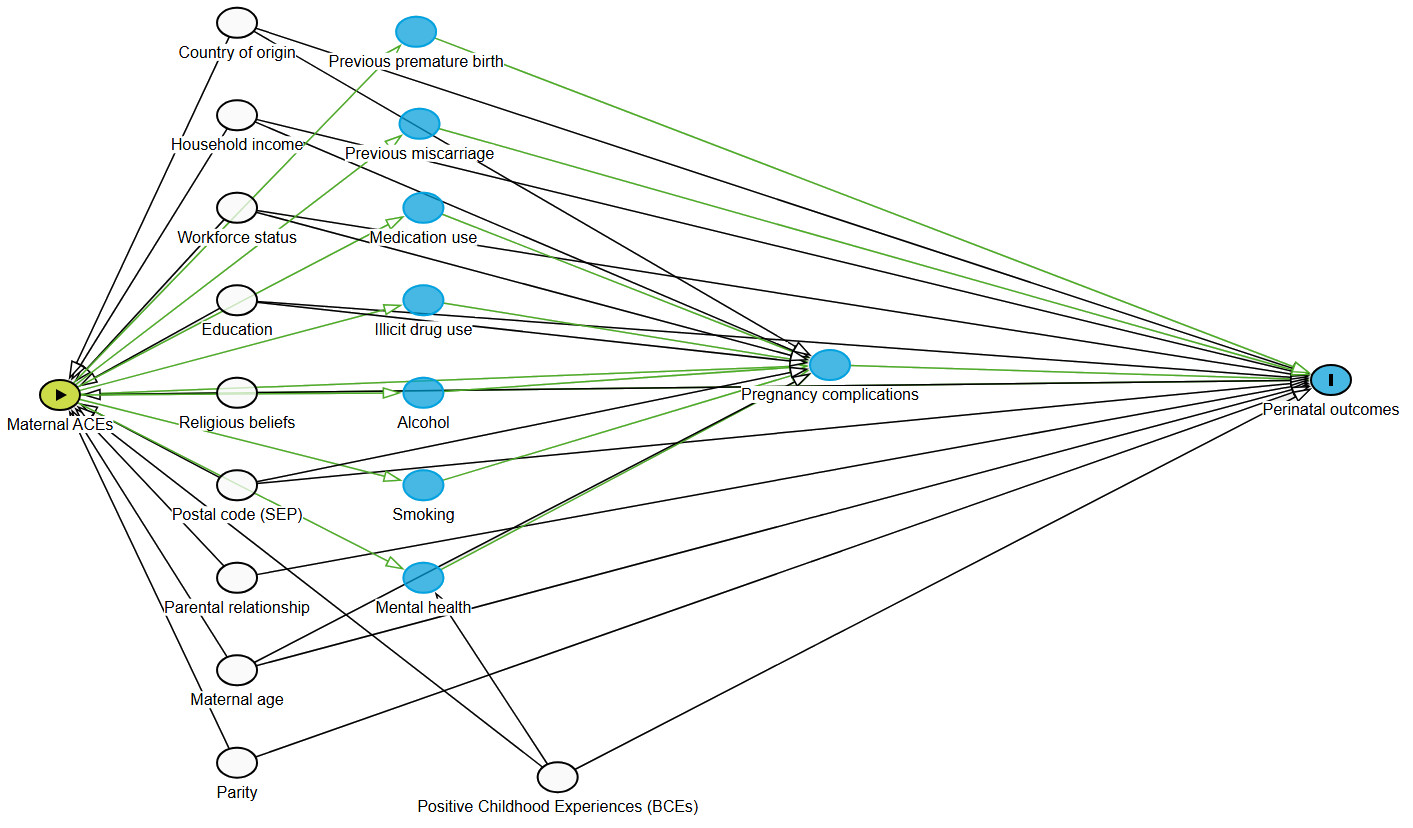

**Supplementary Table 1. Complete model output for unadjusted logistic regressions between ACEs (categorical) and outcomes.**

| **term** | **estimate** | **std.error** | **statistic** | **p.value** | **OR** | **OR_low** | **OR_high** |
| --- | --- | --- | --- | --- | --- | --- | --- |
| Model: premature ~ aces_cat | | |  |  |  |  |  |
| (Intercept) | | 0,23 | -11,16 | 0 | 0,08 | 0,05 | 0,12 |
| aces_catModerate | | 0,3 | 0,43 | 0,669 | 1,14 | 0,63 | 2,06 |
| aces_catHigh | | 0,4 | 0,11 | 0,909 | 1,05 | 0,48 | 2,3 |
| Model: sga ~ aces_cat | | |  |  |  |  |  |
| (Intercept) | | 0,19 | -11,04 | 0 | 0,12 | 0,08 | 0,17 |
| aces_catModerate | | 0,26 | -0,26 | 0,794 | 0,93 | 0,56 | 1,56 |
| aces_catHigh | | 0,32 | 0,82 | 0,413 | 1,3 | 0,7 | 2,42 |
| Model: nicu ~ aces_cat | | |  |  |  |  |  |
| (Intercept) | | 0,25 | -11,22 | 0 | 0,06 | 0,04 | 0,1 |
| aces_catModerate | | 0,33 | 0,04 | 0,964 | 1,01 | 0,53 | 1,95 |
| aces_catHigh | | 0,49 | -0,67 | 0,502 | 0,72 | 0,28 | 1,87 |
| Model: pregnancy complications ~ aces_cat | | | | |  |  |  |
| (Intercept) | | 0,13 | -7,22 | 0 | 0,39 | 0,31 | 0,51 |
| aces_catModerate | | 0,17 | 0,95 | 0,342 | 1,17 | 0,84 | 1,64 |
| aces_catHigh | | 0,21 | 2,43 | 0,015 | 1,68 | 1,11 | 2,56 |
| Model: any outcome ~ aces_cat | | | |  |  |  |  |
| (Intercept) | | 0,12 | -2,94 | 0,003 | 0,7 | 0,56 | 0,89 |
| aces_catModerate | | 0,16 | 0,01 | 0,991 | 1 | 0,73 | 1,37 |
| aces_catHigh | | 0,21 | 1,9 | 0,057 | 1,49 | 0,99 | 2,24 |

**Supplementary Table 2. Complete model output for unadjusted logistic regressions between ACEs (numerical) and outcomes.**

| **term** | **estimate** | **std.error** | **statistic** | **p.value** | **OR** | **OR_low** | **OR_high** |
| --- | --- | --- | --- | --- | --- | --- | --- |
| Model: premature ~ acescore | | |  |  |  |  |  |
| (Intercept) | | 0,19 | -13,72 | 0 | 0,08 | 0,05 | 0,11 |
| acescore |  | 0,07 | 0,61 | 0,545 | 1,04 | 0,91 | 1,19 |
| Model: sga ~ acescore | | |  |  |  |  |  |
| (Intercept) | | 0,16 | -13,48 | 0 | 0,11 | 0,08 | 0,16 |
| acescore |  | 0,06 | 0,48 | 0,633 | 1,03 | 0,91 | 1,16 |
| Model: NICU_opname ~ acescore | | | |  |  |  |  |
| (Intercept) | | 0,2 | -13,35 | 0 | 0,07 | 0,04 | 0,1 |
| acescore |  | 0,09 | -0,69 | 0,49 | 0,94 | 0,79 | 1,12 |
| Model: complicaties ~ acescore | | | |  |  |  |  |
| (Intercept) | | 0,1 | -8,51 | 0 | 0,41 | 0,34 | 0,5 |
| acescore |  | 0,04 | 2,17 | 0,03 | 1,09 | 1,01 | 1,17 |
| Model: anyoutcome ~ acescore | | | |  |  |  |  |
| (Intercept) | | 0,1 | -3,88 | 0 | 0,68 | 0,56 | 0,83 |
| acescore |  | 0,04 | 1,71 | 0,088 | 1,07 | 0,99 | 1,15 |

**Supplementary Table 3. Complete model output for unadjusted linear regressions between ACEs (categorical) and outcomes.**

| **term** | **estimate** | **std.error** | **statistic** | **p.value** | **conf.low** | **conf.high** |
| --- | --- | --- | --- | --- | --- | --- |
| Model: birthweight ~ aces_cat | | | |  |  |  |
| (Intercept) | 3424,82 | 33,31 | 102,83 | 0 | 3359,55 | 3490,1 |
| aces_catModerate | -19,92 | 44,48 | -0,45 | 0,654 | -107,1 | 67,26 |
| aces_catHigh | -24,39 | 58,42 | -0,42 | 0,676 | -138,89 | 90,11 |
| Model: Gestational_age ~ aces_cat | | |  |  |  |  |
| (Intercept) | 39,07 | 0,12 | 316,06 | 0 | 38,83 | 39,31 |
| aces_catModerate | -0,09 | 0,16 | -0,56 | 0,578 | -0,41 | 0,23 |
| aces_catHigh | 0,04 | 0,22 | 0,18 | 0,856 | -0,39 | 0,47 |

**Supplementary Table 4. Complete model output for unadjusted linear regressions between ACEs (numerical) and outcomes.**

| **term** | **estimate** | **std.error** | **statistic** | **p.value** | **conf.low** | **conf.high** |
| --- | --- | --- | --- | --- | --- | --- |
| Model: birthweight ~ acescore | | | |  |  |  |
| (Intercept) | 3436,99 | 27,8 | 123,63 | 0 | 3382,5 | 3491,48 |
| acescore | -10,66 | 10,8 | -0,99 | 0,324 | -31,82 | 10,5 |
| Model: Gestational_age ~ acescore | | |  |  |  |  |
| (Intercept) | 39,04 | 0,1 | 381,78 | 0 | 38,84 | 39,24 |
| acescore | 0 | 0,04 | -0,08 | 0,937 | -0,08 | 0,08 |

**Supplementary Table 5. Complete model output for adjusted logistic regressions between ACEs (categorical) and outcomes.**

| **term** | **OR** | **OR_low** | **OR_high** | **std.error** | **statistic** | **p.value** |  |
| --- | --- | --- | --- | --- | --- | --- | --- |
| Model: prematurity ~ aces_cat + education_bin | | |  |  |  |  |  |
| (Intercept) | 0,07 | 0,04 | 0,11 | 0,24 | -11,16 | 0 |  |
| aces_catModerate | 1,11 | 0,61 | 2,01 | 0,3 | 0,34 | 0,737 |  |
| aces_catHigh | 0,95 | 0,43 | 2,12 | 0,41 | -0,12 | 0,903 |  |
| education_binLower/Middle | 1,6 | 0,83 | 3,09 | 0,34 | 1,39 | 0,165 |  |
| Model: sga_combined ~ aces_cat + education_bin | | |  |  |  |  |  |
| (Intercept) | 0,12 | 0,08 | 0,17 | 0,2 | -10,92 | 0 |  |
| aces_catModerate | 0,93 | 0,56 | 1,56 | 0,26 | -0,27 | 0,786 |  |
| aces_catHigh | 1,28 | 0,68 | 2,42 | 0,32 | 0,77 | 0,441 |  |
| education_binLower/Middle | 1,06 | 0,57 | 1,97 | 0,32 | 0,18 | 0,86 |  |
| Model: NICU_admission ~ aces_cat + education_bin | | |  |  |  |  |  |
| (Intercept) | 0,06 | 0,04 | 0,1 | 0,25 | -11,04 | 0 |  |
| aces_catModerate | 1,01 | 0,53 | 1,95 | 0,33 | 0,04 | 0,966 |  |
| aces_catHigh | 0,72 | 0,27 | 1,89 | 0,49 | -0,67 | 0,504 |  |
| education_binLower/Middle | 1,01 | 0,44 | 2,35 | 0,43 | 0,03 | 0,972 |  |
| Model: pregnancy_complications ~ aces_cat + education_bin | | |  |  |  |  |  |
| (Intercept) | 0,37 | 0,29 | 0,48 | 0,13 | -7,48 | 0 |  |
| aces_catModerate | 1,15 | 0,82 | 1,6 | 0,17 | 0,8 | 0,422 |  |
| **aces_catHigh** | **1,55** | **1,01** | **2,37** | **0,22** | **2** | **0,045** | * |
| education_binLower/Middle | 1,58 | 1,07 | 2,34 | 0,2 | 2,3 | 0,021 |  |
| Model: anyoutcome ~ aces_cat + education_bin | | |  |  |  |  |  |
| (Intercept) | 0,67 | 0,52 | 0,85 | 0,12 | -3,32 | 0,001 |  |
| aces_catModerate | 0,97 | 0,71 | 1,33 | 0,16 | -0,18 | 0,86 |  |
| aces_catHigh | 1,35 | 0,89 | 2,06 | 0,21 | 1,42 | 0,155 |  |
| education_binLower/Middle | 1,68 | 1,13 | 2,49 | 0,2 | 2,59 | 0,01 |  |

**Supplementary Table 6. Complete model output for adjusted logistic regressions between ACEs (numerical) and outcomes.**

| **term** | **OR** | **OR_low** | **OR_high** | **std.error** | **statistic** | **p.value** |  |
| --- | --- | --- | --- | --- | --- | --- | --- |
| Model: prematurity ~ acescore + education_bin | | |  |  |  |  |  |
| (Intercept) | 0,08 | 0,05 | 0,11 | 0,19 | -13,63 | 0 |  |
| acescore | 1,02 | 0,89 | 1,18 | 0,07 | 0,32 | 0,745 |  |
| education_binLower/Middle | 1,5 | 0,75 | 3 | 0,35 | 1,15 | 0,25 |  |
| Model: sga_combined ~ acescore + education_bin | | |  |  |  |  |  |
| (Intercept) | 0,11 | 0,08 | 0,16 | 0,16 | -13,35 | 0 |  |
| acescore | 1,02 | 0,91 | 1,16 | 0,06 | 0,38 | 0,704 |  |
| education_binLower/Middle | 1,13 | 0,59 | 2,16 | 0,33 | 0,36 | 0,722 |  |
| Model: NICU_admission ~ acescore + education_bin | | |  |  |  |  |  |
| (Intercept) | 0,07 | 0,04 | 0,1 | 0,21 | -13,03 | 0 |  |
| acescore | 0,95 | 0,8 | 1,13 | 0,09 | -0,55 | 0,581 |  |
| education_binLower/Middle | 0,74 | 0,28 | 1,94 | 0,5 | -0,62 | 0,537 |  |
| Model: pregnancy_complications ~ acescore + education_bin | | |  |  |  |  |  |
| (Intercept) | 0,4 | 0,32 | 0,49 | 0,11 | -8,67 | 0 |  |
| **acescore** | **1,07** | **0,99** | **1,16** | **0,04** | **1,72** | **0,085** | . |
| education_binLower/Middle | 1,46 | 0,97 | 2,21 | 0,21 | 1,81 | 0,071 |  |
| Model: anyoutcome ~ acescore + education_bin | | |  |  |  |  |  |
| (Intercept) | 0,66 | 0,54 | 0,8 | 0,1 | -4,17 | 0 |  |
| acescore | 1,05 | 0,97 | 1,13 | 0,04 | 1,21 | 0,226 |  |
| education_binLower/Middle | 1,55 | 1,02 | 2,34 | 0,21 | 2,07 | 0,039 |  |

**Supplementary Table 7. Complete model output for adjusted linear regressions between ACEs (categorical) and outcomes.**

| **term** | **estimate** | **std.error** | **statistic** | **p.value** | **conf.low** | **conf.high** |
| --- | --- | --- | --- | --- | --- | --- |
| Model: Gestational_age ~ aces_cat + education_bin | |  |  |  |  |  |
| (Intercept) | 39,12 | 0,13 | 312,7 | 0 | 38,87 | 39,36 |
| aces_catModerate | -0,07 | 0,16 | -0,42 | 0,677 | -0,39 | 0,25 |
| aces_catHigh | 0,12 | 0,22 | 0,56 | 0,577 | -0,31 | 0,55 |
| education_binLower/Middle | -0,45 | 0,21 | -2,19 | 0,029 | -0,86 | -0,05 |
| Model: birthweight ~ aces_cat + education_bin | | | |  |  |  |
| (Intercept) | 3431,82 | 33,8 | 101,54 | 0 | 3365,59 | 3498,06 |
| aces_catModerate | -16,48 | 44,56 | -0,37 | 0,712 | -103,81 | 70,86 |
| aces_catHigh | -12,16 | 59,28 | -0,21 | 0,837 | -128,34 | 104,01 |
| education_binLower/Middle | -67,06 | 55,52 | -1,21 | 0,227 | -175,87 | 41,75 |

Supplementary Table 8. Complete model output for adjusted linear regressions between ACEs (numerical) and outcomes.

| **term** | **estimate** | **std.error** | **statistic** | **p.value** | **conf.low** | **conf.high** |
| --- | --- | --- | --- | --- | --- | --- |
| Model: birthweight ~ acescore + education_bin | | | |  |  |  |
| (Intercept) | 3441,56 | 28,28 | 121,68 | 0 | 3386,12 | 3496,99 |
| acescore | -8,54 | 11,06 | -0,77 | 0,44 | -30,22 | 13,15 |
| education_binLower/Middle | -52,18 | 59,27 | -0,88 | 0,379 | -168,35 | 63,98 |
| Model: Gestational_age ~ acescore + education_bin | |  |  |  |  |  |
| (Intercept) | 39,07 | 0,1 | 376,59 | 0 | 38,87 | 39,27 |
| acescore | 0,01 | 0,04 | 0,29 | 0,775 | -0,07 | 0,09 |
| education_binLower/Middle | -0,36 | 0,22 | -1,64 | 0,101 | -0,79 | 0,07 |
